# Blood transcriptomes of SARS-CoV-2 infected kidney transplant recipients demonstrate immune insufficiency

**DOI:** 10.1101/2022.01.31.22270203

**Authors:** Zeguo Sun, Zhongyang Zhang, Khadija Banu, Yorg Al Azzi, Anand Reghuvaran, Samuel Fredericks, Marina Planoutene, Susan Hartzell, John Pell, Gregory Tietjen, William Asch, Sanjay Kulkarni, Richard Formica, Meenakshi Rana, Weijia Zhang, Enver Akalin, Paolo Cravedi, Peter S. Heeger, Madhav C. Menon

## Abstract

**Background:** Kidney transplant recipients (KTRs) with COVID-19 have poor outcomes compared to non-KTRs. To provide insight into management of immunosuppression during acute illness, we studied immune signatures from the peripheral blood during and after COVID-19 infection from a multicenter KTR cohort.□

**Methods:** Clinical data were collected by chart review. PAXgene blood RNA was poly-A selected and RNA sequencing was performed to evaluate transcriptome changes.

**Results:** A total of 64 cases of COVID-19 in KTRs were enrolled, including 31 acute cases (< 4 weeks from diagnosis) and 33 post-acute cases (>4 weeks). In the blood transcriptome of acute cases, we identified differentially expressed genes (DEGs) in positive or negative association COVID-19 severity scores. Functional enrichment analyses showed upregulation of neutrophil and innate immune pathways, but downregulation of T-cell and adaptive immune-activation pathways proportional to severity score. This finding was independent of lymphocyte count and despite reduction in immunosuppression (IS) in most KTRs. Comparison with post-acute cases showed “normalization” of these enriched pathways after >4 weeks, suggesting recovery of adaptive immune system activation despite reinstitution of IS. The latter analysis was adjusted for COVID-19 severity score and lymphocyte count. DEGs associated with worsening disease severity in a non-KTR cohort with COVID-19 (GSE152418) showed significant overlap with KTRs in these identified enriched pathways.

**Conclusion:** Blood transcriptome of KTRs affected by COVID-19 shows decrease in T-cell and adaptive immune activation pathways during acute disease that associate with severity despite IS reduction and show recovery after acute illness.

**Significance statement:** Kidney transplant recipients (KTRs) are reported to have worse outcomes with COVID-19, and empiric reduction of maintenance immunosuppression is pursued. Surprisingly, reported rates of acute rejection have been low despite reduced immunosuppression. We evaluated the peripheral blood transcriptome of 64 KTRs either during or after acute COVID-19. We identified transcriptomic signatures consistent with suppression of adaptive T-cell responses which significantly associated with disease severity and showed evidence of recovery after acute disease, even after adjustment for lymphocyte number. Our transcriptomic findings of immune-insufficiency during acute COVID-19 provide an explanation for the low rates of acute rejection in KTRs despite reduced immunosuppression. Our data support the approach of temporarily reducing T –cell-directed immunosuppression in KTRs with acute COVID-19.

## Introduction

Coronavirus disease 2019 (COVID-19), caused by Severe Acute Respiratory Syndrome Coronavirus 2 (SARS-CoV-2) infection, is an ongoing global pandemic(1). Wide reporting of cohort studies from multiple continents have established clinical and epidemiologic risk factors for disease severity including advanced age and presence of comorbidities such as cardiovascular disease, diabetes, lung disease, and obesity(2-7).

Immunosuppressed kidney transplant recipients (KTRs) are largely reported to have poor outcomes compared to non-transplant patients (hereafter non-KTRs)(8-11),(12, 13). Since immunosuppression (IS) could inhibit the development of protective anti-COVID-19 immunity, most centers have empirically reduced anti-rejection immunosuppression in KTRs with COVID-19 from the onset of the pandemic. However, severe COVID-19 infection itself has demonstrated significant immune dysregulation(14) manifested as cytokine upregulation(15-17) along with simultaneous HLA class II downregulation on monocytes (18), lymphopenia(19), and T-cell exhaustion (20). Additionally, while we initially observed preserved ability to mount anti-COVID-19 adaptive T-cell responses in KTRs(21), we and others also reported reduced humoral responses in KTRs post COVID-19 infection(22).

Hence understanding immune signatures from the peripheral blood of KTRs during and after COVID-19 infection would provide insight into management of IS during acute disease. Furthermore, evaluating immune signatures after reinstitution of IS following the acute disease episode in KTRs is also relevant to understanding the return of immunocompetence in the setting of the ongoing pandemic and the risk for subsequent graft rejection. We therefore studied peripheral transcriptomes of KTRs with acute (≤4 weeks) and post-acute (>4 weeks) COVID-19 from two major transplant centers in New York City, over the course of the pandemic (8, 11). We identified significant downregulation of T cell-mediated immune activation signatures in KTRs with COVID-19 that were correlated with disease severity, but significant recovery after the acute episode despite reinstitution of the IS regimen. We compared these data with COVID-19 infected non-KTRs to reveal KTR- or IS-unique signatures.

## Results

### Demographics of the study cohort and outcomes

Kidney transplant recipients (KTRs) diagnosed with COVID-19 were enrolled in the study from both centers (Mount Sinai and Montefiore) between 5/1/2020 and 5/30/2021 (Fig 1). A total of 64 KTRs remained in the final dataset for transcriptome analysis, in which 31 KTRs were enrolled within the first 4 weeks of COVID-19 illness or hospitalization (hereafter named acute or early patients), while 33 KTRs were separately enrolled after 4 weeks of illness (late or post-acute patients; see methods). Table 1 lists demographics and clinical characteristics of the study cohort stratified by early and late patients. The majority of KTRs had transplant vintage greater than 2 years post-transplantation at the time of COVID-19 diagnosis. As shown, ∼20% of the KTRs were self-reported African Americans (AAs), with a larger proportion of AAs in early (acute) enrollees. Acute KTRs had slightly larger BMI than post-acute patients (mean 30.4 vs. 27.6 kg/m^2^). The lymphocyte counts in acute KTRs were significantly higher than post-acute KTRs (mean 1.45 vs. 0.80 10^3^/ml). Acute KTRs tended to have higher COVID-19 severity score than post-acute KTRs (p < 0.001). No other significant differences were identified between the two groups in clinical or demographic characteristics. Peak serum creatinine during COVID-19 illness was available in 72% of patients, and acute kidney injury (AKI) during acute illness (>25% change from baseline creatinine) occurred in 87.1% of this group. Eight patients died in the cohort, and as expected these were in acute COVID KTR group. No cases of biopsy proven acute rejection were reported in this group. Three patients lost their grafts over the duration of the study period, 1 in acute group and 2 in post-acute group with no statistically significant difference.

**Figure 1.**
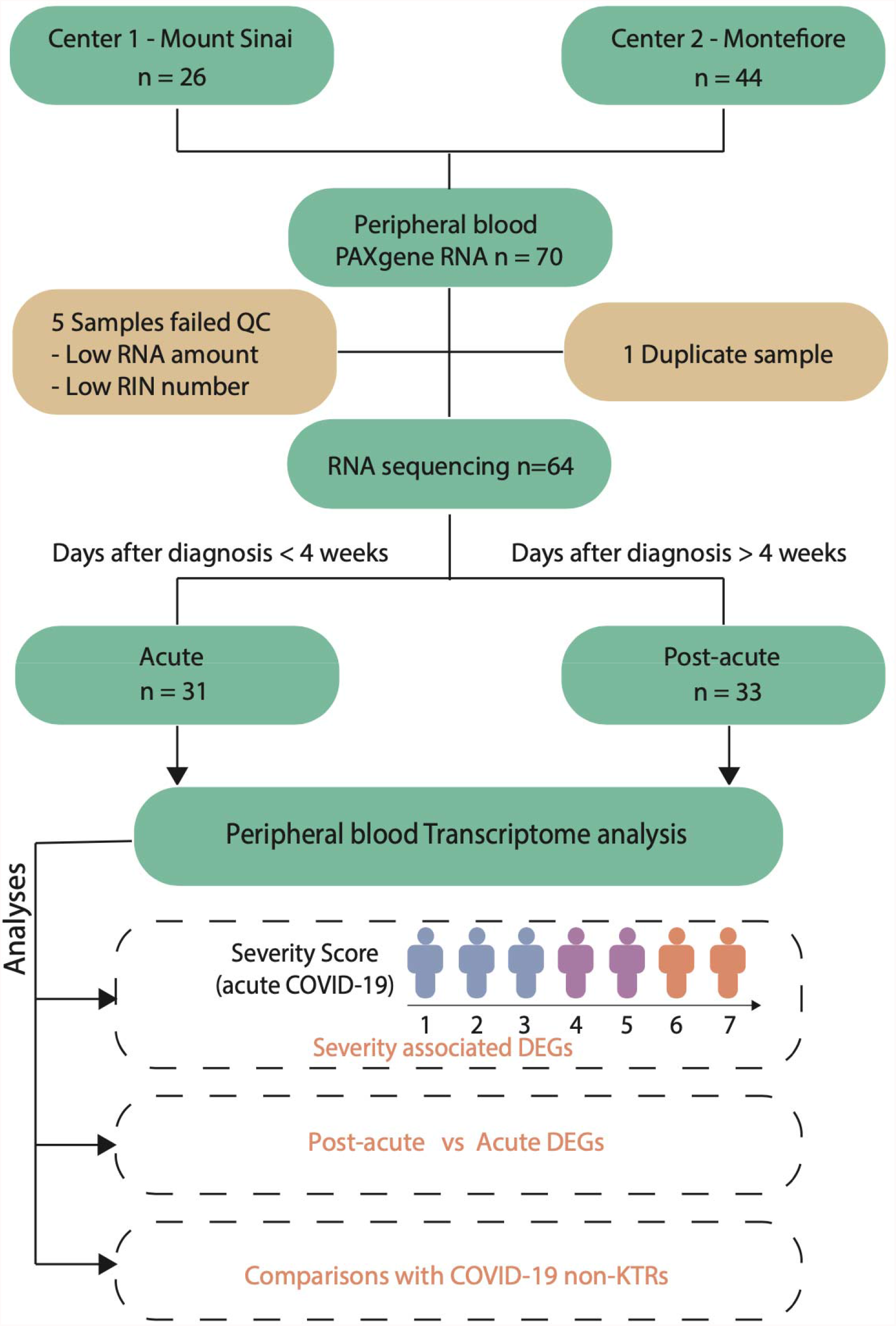
Flow diagram of study design. A total of 70 KTRs with documented evidence of SARS-CoV-2 infection were enrolled from two transplant centers in New York. Blood was collected (∼2.5ml) in a PAXgene tube for RNA extraction and transcriptome quantification. After quality control (QC), a total of 64 KTR transcriptomes remained for RNA sequencing (n = 31 acute cases and n = 33 post-acute cases). One patient in post-acute group was diagnosed by antecedent COVID-19 IgG testing. The severity scores of included patients during acute COVID-19 illness was evaluated (WHO severity score ranging from 1 to 7 indicating escalating severity). Significantly differentially expressed genes (DEGs) were identified from the blood transcriptomes of enrollees in the following analyses – DEGs associated ordinally with severity score, and DEGs between post-acute and acute patients. External non-KTR datasets were downloaded to test validation of findings.

**Table 1.**
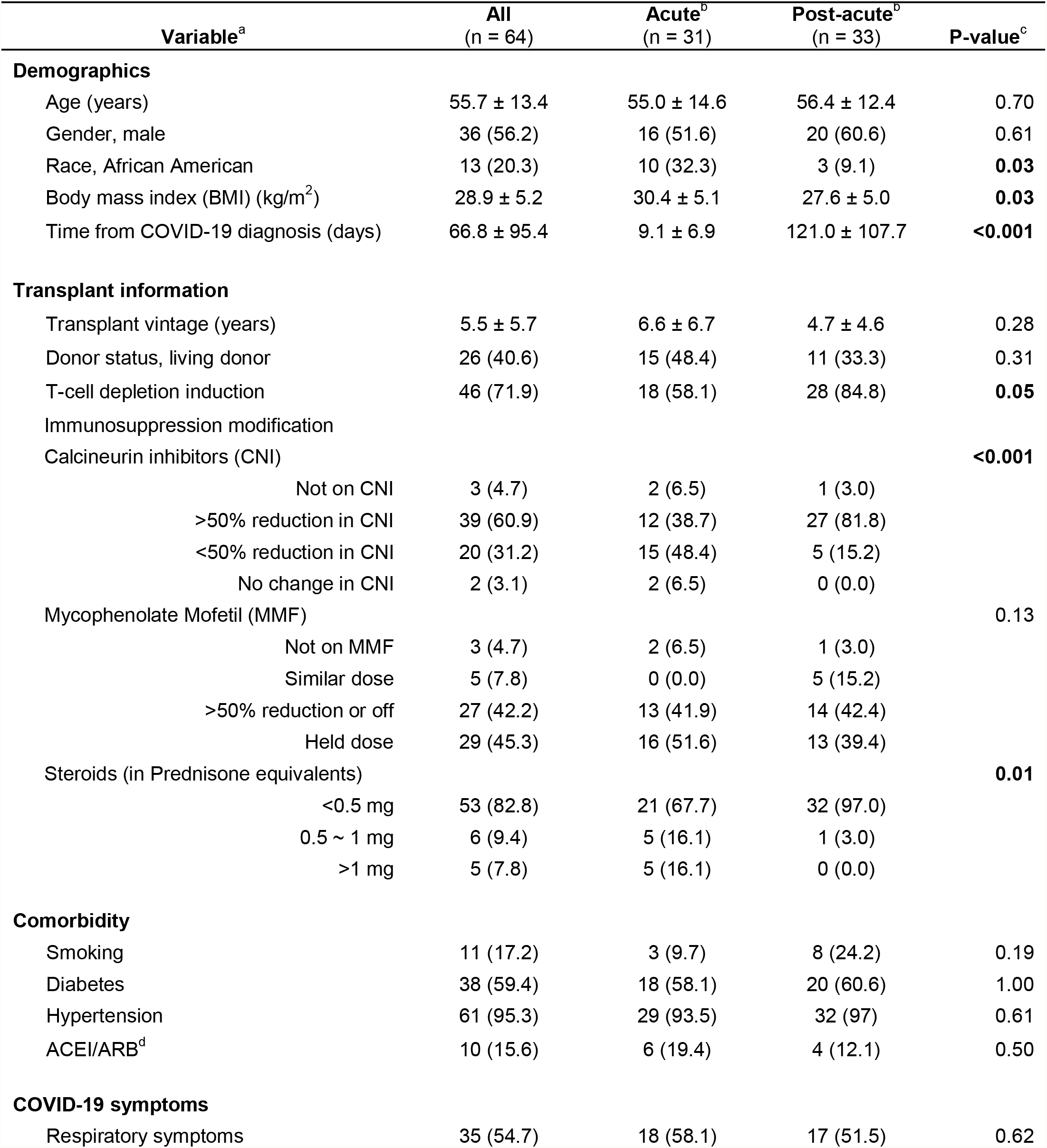
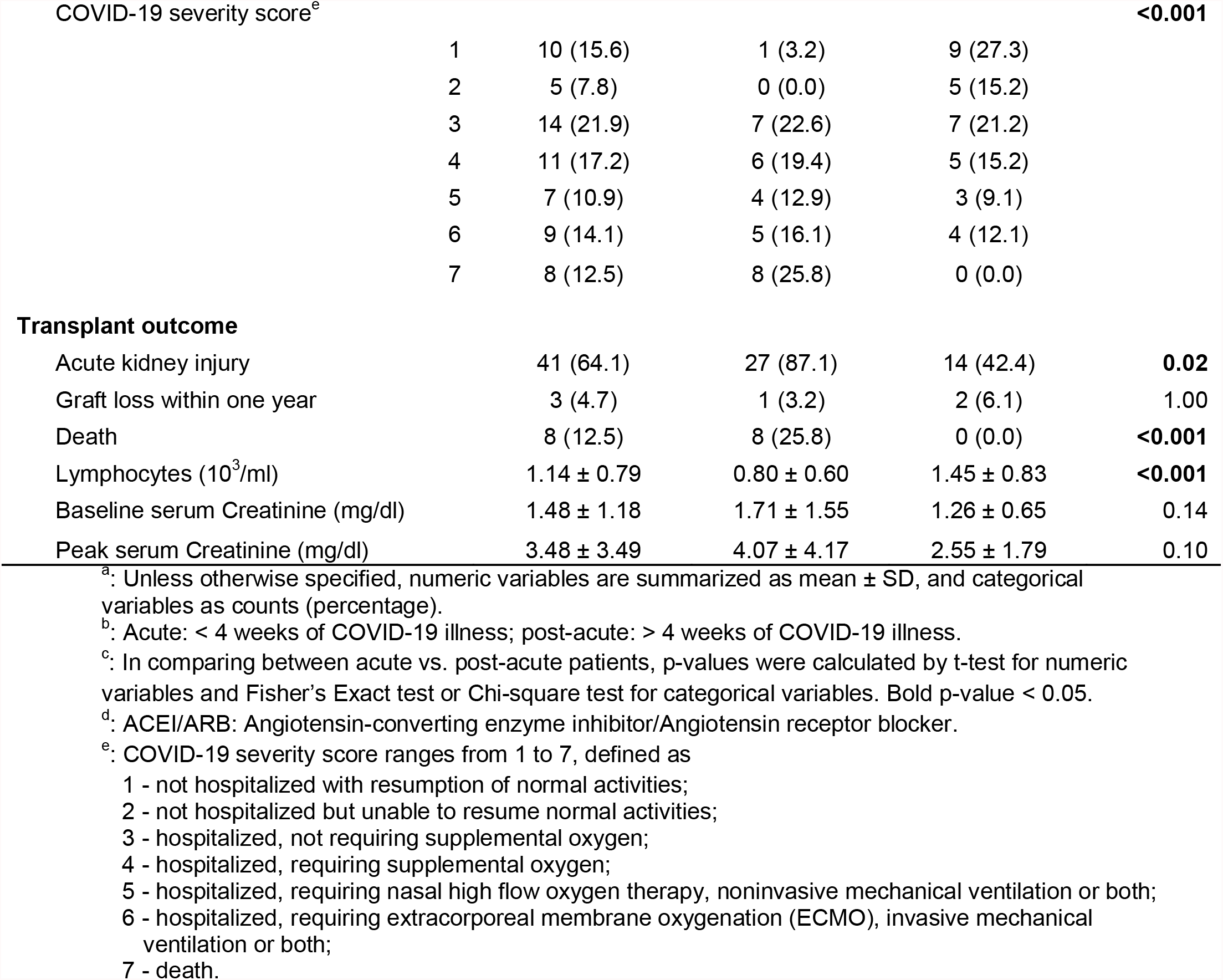
Demographic and clinicopathologic characteristics of the study cohort.

### Transcriptome in KTRs during acute COVID-19 episode shows signature of neutrophil activation and deficient adaptive immune responses associated with increasing severity

We first focused on KTRs with acute COVID-19 to examine peripheral blood transcriptomic changes as a function of disease severity score. Table S1 summarizes clinical and demographic characteristics of acute KTRs stratified by the severity score. Within strata of increased severity, the patients were older, tended to have more respiratory symptoms, and were on significantly higher dose of steroid therapy. As expected, Mycophenolate Mofetil (MMF) therapy was reduced or withheld in more than half of all acute patients, and Calcineurin inhibitors (CNI) were continued at the same dose only in 25% of mild cases (Supplementary Table S1). AKI during acute episode (>25% change in serum creatinine from baseline) was universally seen in this cohort, but baseline chronic kidney disease in allograft (by higher baseline creatinine) was more common within the intermediate severity stratum. As reported before (23, 24), association of lymphopenia with disease severity was also observed in our cohort.

We then correlated changes in peripheral transcriptomes with increasing severity scores. Genes with expression levels that associated with increasing severity score (treated as numeric variable in linear regression model) were identified at nominal p-value ≤ 0.01 (see methods). Differentially expressed genes (DEGs) in positive association (n = 325) and negative association (n = 216) with the severity score are shown in Fig 2A. Since lymphopenia correlated with severity score in the KTR cohort [Spearman R=-0.36; P<0.01; Fig S1], these analyses were adjusted for clinical variables (see methods), including simultaneous lymphocyte count. To explore pathways enriched in the severity correlated DEGs, EnrichR was used with pathways (gene sets) defined from multiple databases (25) (Fig 2B and Supplementary Table S2). Neutrophil activation, neutrophil degranulation, cytokine release and signaling (TNFα-NFκB) were all upregulated with increasing severity score. In contrast, adaptive immune system pathways (Th1, Th2, and Th17 cell differentiation, antigen processing and presentation, and IFNγ signaling) were downregulated as severity scores increased. To investigate whether the enriched downregulation signals in T-cell activation pathways reflected absolute T-cell count depletion during severe COVID-19, we further examined a subset of acute KTRs with CD3-positive T-cell counts generated from flow cytometry analysis (n=16/31; see Methods). As expected, the T-cell count was correlated with lymphocyte count; however it did not correlate with the severity score in this smaller subset of acute KTRs (Supplementary Figure S1). In this subset, when adjusted for T-cell count in the association analysis with severity, neutrophilic and T-cell activation pathways remained enriched (as in the whole acute cohort), albeit at lower significance (Supplementary Fig S2). Gene set enrichment analysis (GSEA)(26) of representative pathways (neutrophil degranulation and antigen processing and presentation) highlighted the regulation direction of top-ranked transcripts in association with increasing disease severity (Fig 2C). In the two representative pathways, the trend (increase or decrease) in changes of expression levels with severity scores for top-ranked DEGs are shown in Fig 2D. Serum cytokines such as TNF, IL6, and IL1, which were associated with severity in prior data(15), did not demonstrate corresponding mRNA levels that were proportional to severity score in the peripheral transcriptome. Overall, these data suggest enhanced neutrophil activation with cytokine signaling, and downregulated adaptive and T-cell responses with increasing severity of COVID-19 in KTRs during acute episode.

**Figure 2.**
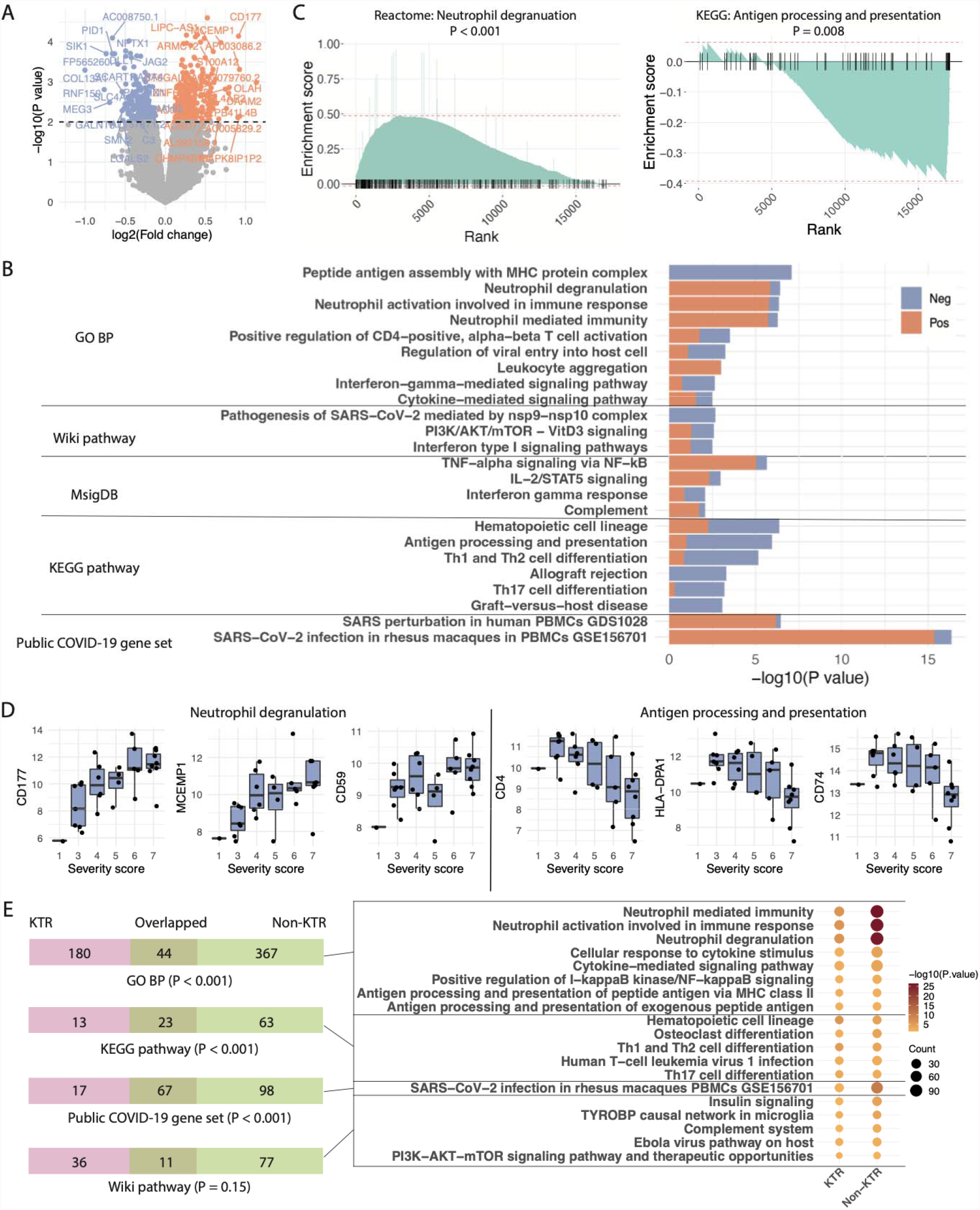
Transcriptome in KTRs with acute COVID-19 shows signature of neutrophil activation and deficient adaptive immune responses with increasing severity. (**A**) Volcano plot shows gene expression change associated with increasing severity score. The log2(fold change) reflects the direction and magnitude of change in expression per unit increase in severity score (see definition in Table 1). Differentially expressed genes (DEGs) were identified with nominal p ≤ 0.01, with positively associated genes colored in red and negatively associated genes in blue. (**B**) Pathways enriched in DEGs with the proportion of positively and negatively associated genes overlapping each pathway colored in red and blue correspondingly. The pathways defined from different sources are labeled on the left and distinguished by horizontal lines. GO BP: Gene Ontology Biological Process. (**C**) Gene Set Enrichment Analysis (GSEA) rank plot of representative pathways, in which genes were ranked by their log2(fold change) from positive to negative association with increasing severity score. The short ticks along the horizontal line at 0 indicate the rank of the genes in the corresponding pathways. (**D**) The trend in expression change of representative genes with an increasing severity score for the neutrophil degranulation and antigen processing and presentation pathways. The scale of expression value is on log2(CPM + 1). CPM: Counts per million reads. (**E**) On the left panel, each Venn diagram (displayed in the form of bars) shows the overlap of enriched pathways of DEGs associated with COVID-19 severity in KTRs and non-KTRs. The overlap significance was evaluated with a hypergeometric test. On the right panel, the representative, overlapped pathways were listed in the dot plot with the size of the dot indicating the number of DEGs in each pathway and the color indicating the enrichment significance.

To compare the blood transcriptomic signatures in COVID-19 infected non-KTRs, we evaluated peripheral transcriptomes from a published cohort of non-KTRs with acute COVID-19 disease and simultaneous reporting of severity of illness (GEO accession: GSE152418)(27). DEGs associated with increasing severity among non-KTRs were identified at nominal p-value ≤ 0.01 (898 up-regulated and 636 down-regulated genes). Similar enrichment analysis platforms (as in KTRs) showed significant overlap between KTRs and non-KTRs in identified pathways associated ordinally with severity (Hypergeometric test p-values < 0.05) (Fig 2E). Some representative immune pathways seen in KTR data also consistently appeared in non-KTR data (Fig 2E; Supplementary Table S3). Hence these data suggest downregulation of T-cell and adaptive response-related genes during acute COVID-19 in KTRs and non-KTRs proportional to increasing severity, regardless of immunosuppressant use.

### Transcriptome in post-acute versus acute disease suggests recovery of adaptive immune responses after acute episode

Next, we evaluated clinical characteristics and peripheral transcriptomes of COVID-19 infected KTRs after the acute phase of illness (>4 weeks; mean 121 days). The clinical and demographic variables of acute and post-acute cases are compared in Table 1, and described above. Steroid therapy during the acute phase of illness was more intense in the acute group and likely related to higher severity scores in this group. Post-acute group had lower BMIs and had lower rates of AKI documented during acute disease.

In peripheral transcriptome comparisons between post-acute and acute KTRs, DEGs significantly up- or down-regulated (310 up-regulated and 328 down-regulated genes respectively identified at nominal p ≤ 0.01) are shown in Fig 3A. In these analyses, we adjusted for clinical variables, simultaneous lymphocyte count, and the severity score during acute episode. In contrast to our observation in the acute covid illness cohort, enrichment of pathways related to T-cell activation was seen among *up-regulated* genes in post-acute cases versus acute cases (Fig 3B and C; Supplementary Table S4).

**Figure 3.**
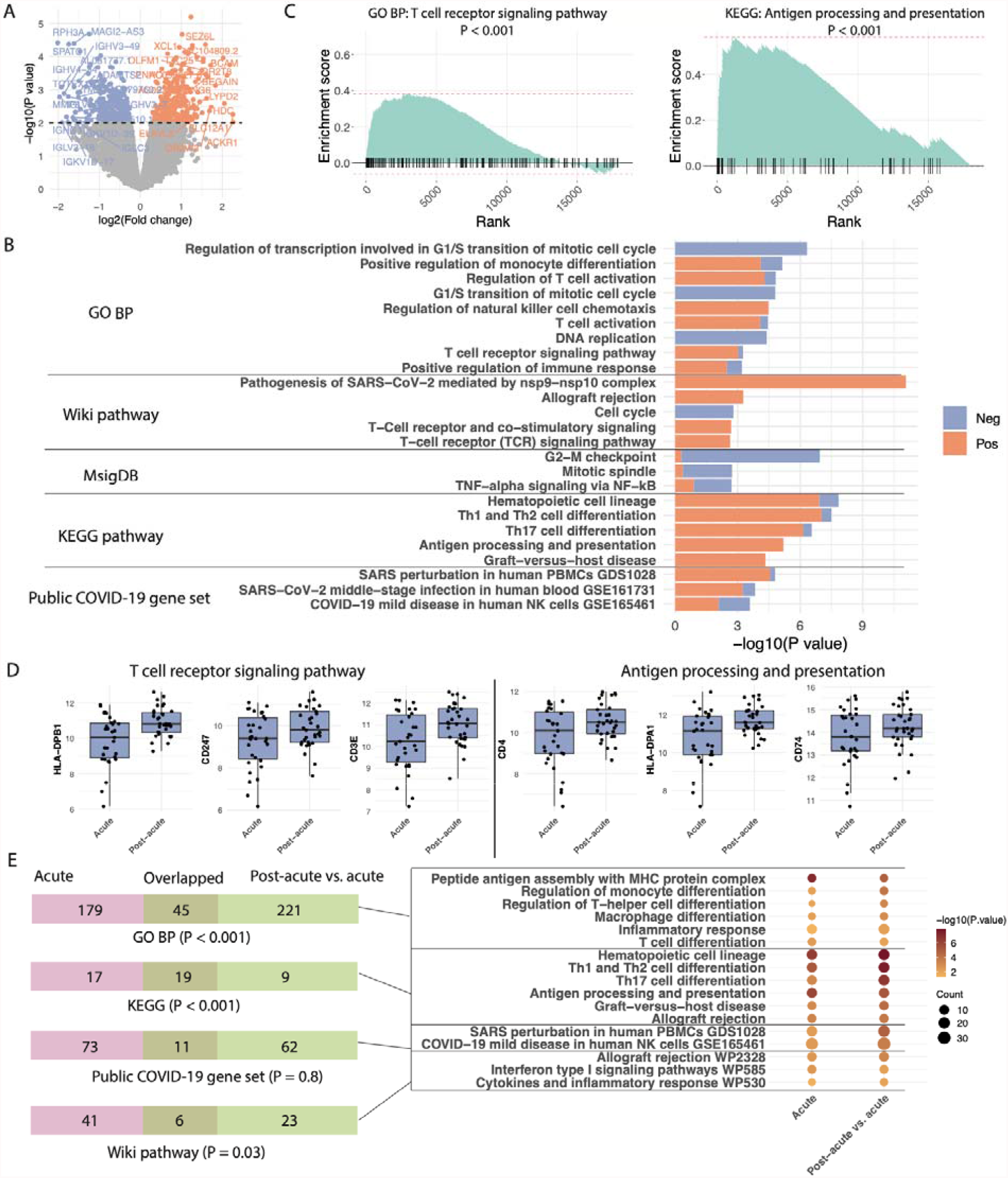
Transcriptome in post-acute versus acute KTRs suggests recovery of adaptive immune responses after acute disease. (**A**) Volcano plot shows gene expression change in post-acute patients as compared to acute patients. DEGs were identified with nominal p ≤ 0.01, with up-regulated genes colored in red and down-regulated genes in blue. (**B**) Pathways enriched in DEGs with the proportion of up- and down-regulated genes colored in red and blue correspondingly. The pathways defined from different sources are labeled on the left and distinguished by horizontal lines. GO BP: Gene Ontology Biological Process. (**C**) GSEA rank plot of representative pathways, in which genes were ranked by their log2(fold change) from up- to down-differential expression. The short ticks along the horizontal line at 0 indicate the rank of the genes in the corresponding pathways. (**D**) Expression change of representative genes in post-acute patients as compared to acute patients. The scale of expression value is on log2(CPM + 1). CPM: Counts per million reads. (**E**) On the left panel, each Venn diagram (displayed in the form of bars) shows the overlap of pathways enriched in DEGs associated with severity score in acute patients and pathways enriched in DEGs in post-acute patients as compared to acute patients. The overlap significance was evaluated with a hypergeometric test. On the right panel, the representative overlapped pathways were listed in the dot plot with the size of the dot indicating the number of DEGs in each pathway and the color indicating the enrichment significance.

Representative DEGS within T-cell activation pathways which showed reduced expression during acute severe disease, here showed significantly increased expression in the post-acute samples (Fig 3D vs Fig 2D). Pathways related to T-cell activation were up-regulated in post-acute cases and showed significant overlap with signaling pathways identified as inversely associated with the severity score (Fig 3E; Supplementary Table S5). This observation suggests a return of adaptive- and T-cell-responses in post-acute cases. In contrast to these distinct changes in the T-cell activation transcriptomes, we did not identify a significant enrichment of neutrophilic activation/degranulation signals in post-acute vs acute comparisons (Supplementary Table S6). Furthermore, *within* post-acute KTRs, neither neutrophilic signaling nor T-cell activation pathways were differentially enriched or associated with COVID-19 severity scores (obtained during the acute COVID-19 episode). These data suggested the return of acute transcriptomic changes to baseline in post-acute cases (Supplementary Figure S3).

To compare these analyses with non-KTR COVID-19 cases where post-acute (convalescent) peripheral transcriptome was reported, we utilized a published RNA-seq dataset (GEO accession: GSE157859)(28). In this dataset, we used longitudinal samples from the same patients (n = 13) obtained within 4 weeks and after 4 weeks of COVID-19 infection. DEGs between post-acute and acute samples in the same patients were identified with the same method, followed by pathway enrichment analyses. The overlapped pathways between KTRs and non-KTRs are illustrated in Supplementary Fig S4 and Table S7. While DEGS within T-cell activation pathways were still identifiable within non-KTRs, significant pathway overlap was not identified between KTRs vs non-KTRs (adjusted for severity in both cases) for most enrichment databases (Fig S4). These data suggest that recovery of immune activation was more detectable in KTRs, possibly due to more pronounced IS during acute COVID-19. Taken together, our data support the conclusion that suppression of T-cell and adaptive immune activation occurs during acute COVID-19 in KTRs, this effect associates with severe disease, and the observed changes recover after acute disease.

## Discussion

Here we studied immunosuppressed kidney transplant recipients from two hospitals at the forefront of the initial COVID-19 pandemic. Based on the reported increased severity of COVID-19 in transplant recipients (8-11), we specifically sought to understand immune signatures in the peripheral blood of KTRs. We attempted to provide an explanation regarding the reported immune dysregulation (14) with marked cytokine upregulation (15-17) in severe COVID-19, particularly in KTRs. As current practice generally involves reducing immunosuppression during COVID-19 in KTRs, we sought evidence to support the safety of this empiric intervention. Our analyses showed significant down-regulation of peripheral blood T cell-mediated immune activation signatures in the majority of the KTRs with acute COVID-19, and that the down regulation correlated with disease severity. Together, these findings are consistent with SARS-CoV-2-induced impairment in adaptive immunity despite IS reduction. These signatures showed overlap with a public dataset of non-KTRs with COVID-19, suggesting some generalizability. Despite reinstitution of the IS regimen in the majority of our post-acute cohort, immune activation pathways appeared to be up-regulated as compared to acute cases, a result that may indicate recovery of T-cell function after the acute episode.

Ours is the first series evaluating the blood transcriptome of KTRs with COVID-19. Our novel findings of evidence of suppression of the T cell/adaptive immune responses in the peripheral transcriptome accompanying COVID-19 and its association with the disease severity have important therapeutic implications for KTRs. While multiple uncontrolled reports have suggested a worse outcome of acute COVID-19 in hospitalized KTRs (8-11), other large datasets did not find this association (13). Further, given the cytokine-mediated immune dysregulation and inflammatory response syndrome that accompanied severe COVID-19 and predicted survival (29-31), it was unclear whether the reflexive practice of reduction/withdrawal of IS in KTRs with COVID-19 is warranted. A large consortium of COVID-19 in transplant recipients concluded that comorbidities contributed to mortality rather than the IS regimen itself (32). Notably, in this multicenter cohort, acute rejection rates were low in COVID-19 KTRs. In these and other data, lymphopenia was an independent predictor of COVID-19 mortality. Hence our blood transcriptomic findings of T cell/adaptive immune suppression during acute COVID-19, proportionate to disease severity after adjustment for lymphocyte count, are likely relevant and informative to therapy. Furthermore, the overlapping signatures in non-KTRs with COVID-19, and the “recovery” of immune signatures specifically in KTRs in the post-acute phase, where IS has been mostly reintroduced (even after adjustment for the severity score), suggest immunosuppression is related to COVID-19 itself, and possibly accentuated by IS in KTRs.

The overlap of signatures of immunosuppression between KTRs and non-KTRs with severe disease is intriguing, and necessitates understanding immune cell dysfunction during COVID-19(33). Multiple non-KTR studies have shown immune cell dysregulation in severe COVID-19, over and above changes in T-lymphocyte numbers (34, 35). While CD4^+^ T cell depletion was observed in severe disease in a large dataset (36), transcriptomic changes associated with severity, were also observed within multiple lineages in single-cell RNA-seq data. Some non-KTR cohorts reported CD8^+^ T cell expansion (not depletion) with severe COVID-19 (37), while others observed heterogenous CD8^+^ T cell numbers, with expansion of some subsets (CD45RA^-^CD27^-^CCR7+ and CD45RA^+^CD27^-^CCR7^-^), but not of other subsets (38). CD8^+^ T cell exhaustion marker (PD1) was reported as increased in severe disease, along with transcriptomic signatures suggestive of a lack of CD4^+^ T cell help, again reflecting CD8^+^ T cell dysfunction (39). Innate immune dysregulation with neutrophilic activation and monocytic dysfunction (with reduced CD4 and HLA-DR expression as well as lack of IL6 and IL1B upregulation as we observed here) was also reported(40). Severe COVID-19 cases showed an early elevation in serum TGFβ, associated with a TGFβ response transcriptional signature in NK cells, leading to NK cell dysfunction with inability to gain early control over the virus (41). Recent evaluations of *ex-vivo* isolated SARS-CoV-2 specific CD4^+^ T cells from severe COVID-19 cases showed expanded albeit dysfunctional virus-specific T-follicular helper cells (with higher ZBED2, TIGIT, and LAG3 expression), as well as Th1 clusters with lower IFNG/IL2 expression (42). In these same data, lower virus specific Th17 cell clusters were observed with severe disease, similar to our transcriptomic data. In addition to these associative data, direct immunomodulatory roles for viral sub-genomic mRNAs have also been proposed (43). Hence, in context of these data showing immune cell dysfunction in acute COVID-19, our findings make a case for reducing or withdrawing transplant-specific T-cell directed IS agents in KTRs (or other immunosuppressed patients) during acute COVID-19 episode.

We acknowledge the lack of serial transcriptomic data from the same patients as a limitation of our work. However, our patients were enrolled during the early peak of the pandemic where in-person visits were minimized, making follow-up sample collection challenging. While we did not enroll a simultaneous non-KTR cohort, we used two different public non-KTR datasets to test the generalizability of our findings. We acknowledge that robust causal inferences cannot be made in our cross-sectional dataset. While overall signals of reduced T cell immunity during acute disease with subsequent recovery later were observed in our data, the signatures among individuals showed some heterogeneity. Our ability to molecularly detect signals consistent with reduced adaptive immunity could be exploited (and applied in external datasets) to develop and test biomarkers to potentially identify individual patients who would best tolerate IS reduction, and possibly guide timing of IS reinstitution.

In summary, we demonstrate that blood transcriptomes of acute COVID-19 in KTRs suggest immune-insufficiency that is proportional to disease severity despite IS reduction. These signatures showed recovery after acute illness. These findings have implications for the pathogenesis and management, including IS reduction or antiviral approaches in COVID-19 infected KTRs.

## Data Availability

All data produced are available online at GSE195796

https://www.ncbi.nlm.nih.gov/geo/query/acc.cgi?acc=GSE195796

## Acknowledgements

Authors acknowledge support from NIH NIAID via ancillary mechanistic grant associated with Grant 3U01AI063594-17S1 awarded to MCM and PC. (Parent award 3U01AI063594 to PSH). MCM also acknowledges research support from NIH RO1DK122164. We acknowledge Ricarda Tomlin at Yale, Brandy Haydel at Mount Sinai and our cohort of patients for participation in this study during the pandemic.This work was supported in part through the computational resources and staff expertise provided by Scientific Computing at the Icahn School of Medicine at Mount Sinai.

## Methods

### Study cohort: COVID-19 infected kidney transplant recipients (KTRs)

***<u>I</u>****<u>nclusion criteria:</u>* Adult KTRs (>18 years), who provided informed consent for sample collection, at both hospital systems (Montefiore Medical Center and Icahn School of Medicine at Mount Sinai) were enrolled during the study period (May 2020 to May 2021). Consented KTRs from inpatient facilities or outpatient transplant clinics were included based on: (a) COVID-19 PCR positivity. Nasopharyngeal and/or oropharyngeal swabs were collected in 3ml viral transport media. RNA extraction and real-time PCR was performed using one of three commercial methods at both institutions; (b) diagnosis of antecedent COVID-19 infection by positive COVID-19 Spike and Nucleocapsid IgG serology. IgG antibody testing was performed using the Abbott SARS-CoV-2 IgG antibody test on the Abbott Architect Immunoassay Analyzer. Testing was performed on serum samples following manufacturer’s instructions. The assay is a chemiluminescent microparticle immunoassay intended for the qualitative detection of IgG antibodies to SARS-CoV-2. COVID-19 infected KTRs were classified as acute (or early) cases (enrollment date <4 weeks from PCR positivity and post-acute (or late) cases (enrollment date >4 weeks from initial COVID-19 diagnosis), with the latter no matter enrolled inpatient or outpatient. We used a cut-off of 4-weeks since the median duration of hospitalization in our- and prior-data(11) were near 14 days. *<u>Exclusion criteria</u>*: pregnant women, inability to give consent, active malignancy, or prior COVID vaccination.

*<u>Covariates:</u>* Basic demographics, transplant-specific variables, important laboratory data during acute phase and follow-up, were obtained by chart review. *<u>Outcomes:</u>* The COVID-19 severity scores during acute phase were categorized in 7 ordinal groups as reported before (21) and was regarded as the primary outcome. Secondary outcomes included need for ICU care (severity score = 6 and 7), AKI, and death. Graft loss within one year and death after enrollment were obtained by chart review of electronic medical records.

### Sample collection

A single sample of blood (2.5-5ml) was collected for peripheral blood transcriptome analysis after consent in a PAXgene tube with RNA stabilization reagents. The date of sample collection was considered enrollment date. PAXgene samples from all patients were transported to the central laboratory at Mount Sinai and stored at -80°C. RNA extraction and processing were done in batches of ten by 2 experienced operators. Total RNA was stored at -80°C till sequencing.

### RNA extraction, library preparation and sequencing

QIAamp RNA Blood Kit was used to extract high quality RNA from total blood (<1% DNA contamination; Qiagen, CA, USA). Total RNA (>500 ng per sample; 260/280 ratio = 2.07 ± 0.08) was poly-A selected and amplified for library preparation and subsequent sequencing (Genewiz Inc, NJ, USA). 150bp paired-end sequencing was performed on the prepared libraries (Illumina, CA, USA) to generate 20-30 million reads per sample, and raw reads data was stored in FASTQ files for analysis.

### RNA sequencing (RNA-seq) data analysis

Quality of raw sequencing reads were summarized with fastqc v0.11.9(44). After trimming the adapter sequences using cutadapt v2.9 (45), cleaned reads were mapped to the human genome (GRCh38) using STAR 2.5.3a (46) with default parameters. Reads mapped to each gene locus was counted by HTseq 0.11.2 with “-m union --nonunique none”(47).

The gene expression in the form of raw read counts were preprocessed with the voom method in limma (v3.38.3) (48) for subsequent transcriptome analysis. Genes with mean expression value (counts per million) less than 1 across all the samples were excluded from downstream analysis. The association of gene expression with the severity score (treated as numeric variable) was analyzed with limma, adjusted by age, gender, and lymphocyte count. Differential expression analysis between acute and post-acute samples was also performed by limma, adjusted by age, gender, lymphocyte count, and the severity score. Differentially expressed genes (DEGs) were identified at nominal p-value ≤ 0.01, followed by functional enrichment analysis of identified DEGs with EnrichR (v2.1) (25). Enrichment signal of representative pathways were further confirmed by Gene Set Enrichment Analysis (GSEA) (26).

Gene expression profiles from two public datasets (GEO accession: GSE152418 and GSE157859) were downloaded from the GEO database. DEG analysis was performed using the same pipeline as described above. Similar nominal P-value thresholds were set. The information about severity group and time after positive PCR was retrieved from the original publications involving these datasets and from GEO (27, 28).

### Flow cytometry analysis

Flow cytometric analysis was performed on total blood or Ficoll-isolated PBMCs in COVID-19 infected kidney transplant patients as detailed previously.(21) From this dataset, we utilized the data from previously designed multicolor flow cytometry panel to quantify subsets of CD3^+^, CD4^+^, and CD8^+^ T cells in the patients overlapped with the acute KTR cohort of our study (Supplemental Table S8). The following fluorochrome-conjugated anti-human antibodies were used from BD Biosciences: CD3-FITC, CD3-PerCP-Cy5.5, and CD4-APC.

### Statistical analyses

Descriptive statistics (means and standard deviation [SD] for numeric variables; sample size and percentage for categorical variables in respective sample group) were used to summarize the baseline characteristics of the acute and post-acute cases. Categorical variables were compared between different groups (acute vs. post-acute and low, median, and high severity of COVID-19) using the Chi-squared test or Fisher’s exact test, while numeric variables were compared between groups using Student’s t-test (two groups) or ANOVA (three groups). Unless otherwise specified, a p-value < 0.05 indicates statistical significance.

### Study approval

Independent institutional review board (IRB) approvals from Icahn school of medicine at Mount Sinai (IRB 20-03454) and Montefiore Medical Center (IRB 2020-11662) were obtained for prospective sample and data collection before initiation of the study protocol. Informed consent was obtained from participants and/or representative proxies per these protocols.

