## Supplementary figures and tables for "Blood transcriptomes of SARS-CoV-2 infected kidney transplant recipients demonstrate immune insufficiency"

**Supplementary Materials:**

**Transcriptomes of COVID-19 infected kidney transplant recipients demonstrate immune insufficiency and persistent circulating SARS-CoV-2 Spike gene expression**

**Supplementary Figures**

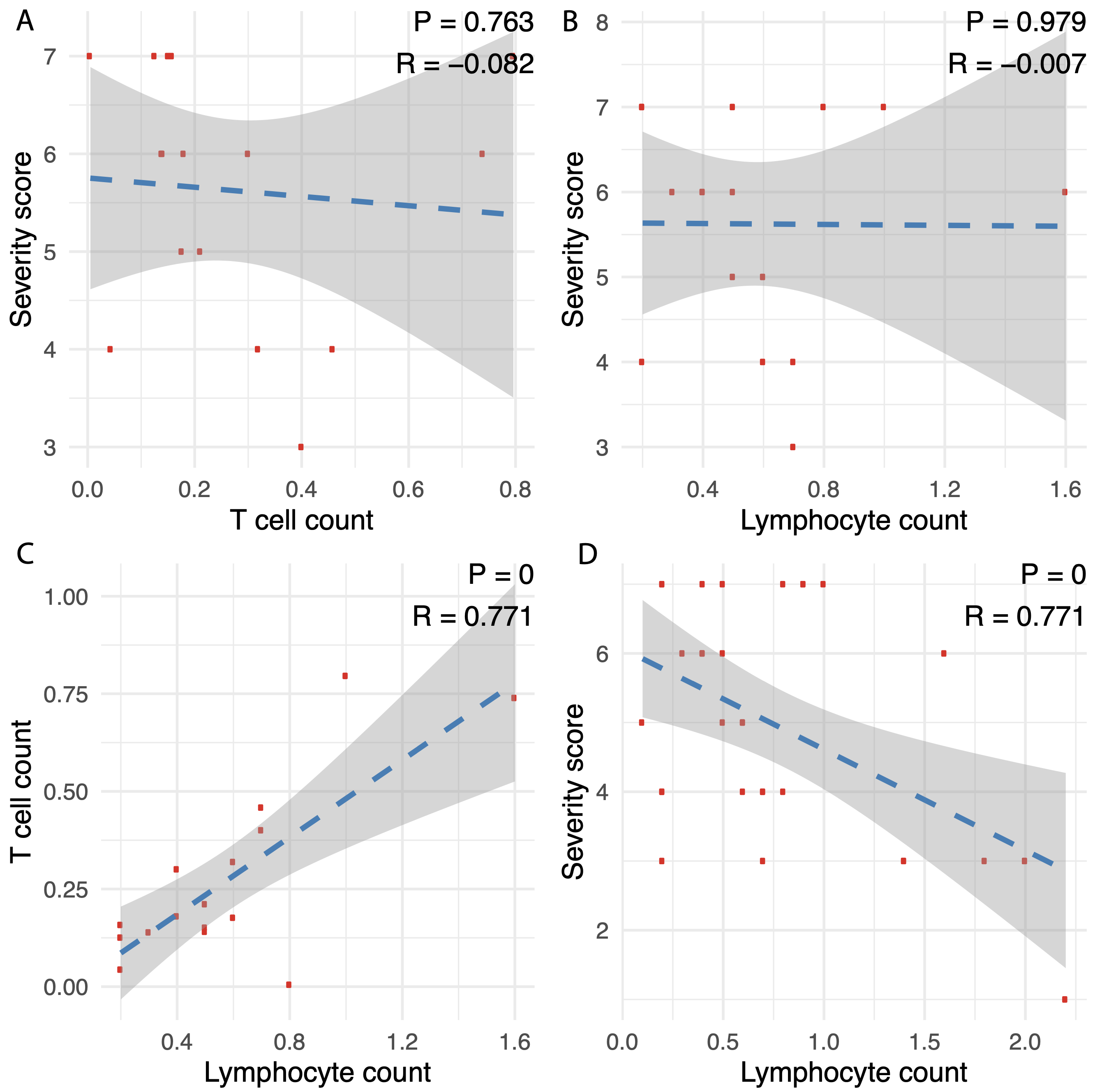

**Figure S1. Correlation of Lymphocyte count, T cell count, and severity score.** Correlation analyses were performed in the 16 acute KTRs with T cell count information for (**A**) T cell count vs. severity score, (**B**) lymphocyte count vs. severity score, and (**C**) lympocypte count vs. T cell count. Pearson correlation coefficient and accompanied p-value are shown. (**D**) correlation between lymphocyte count and severity score was calculated in all 31 acute KTRs.

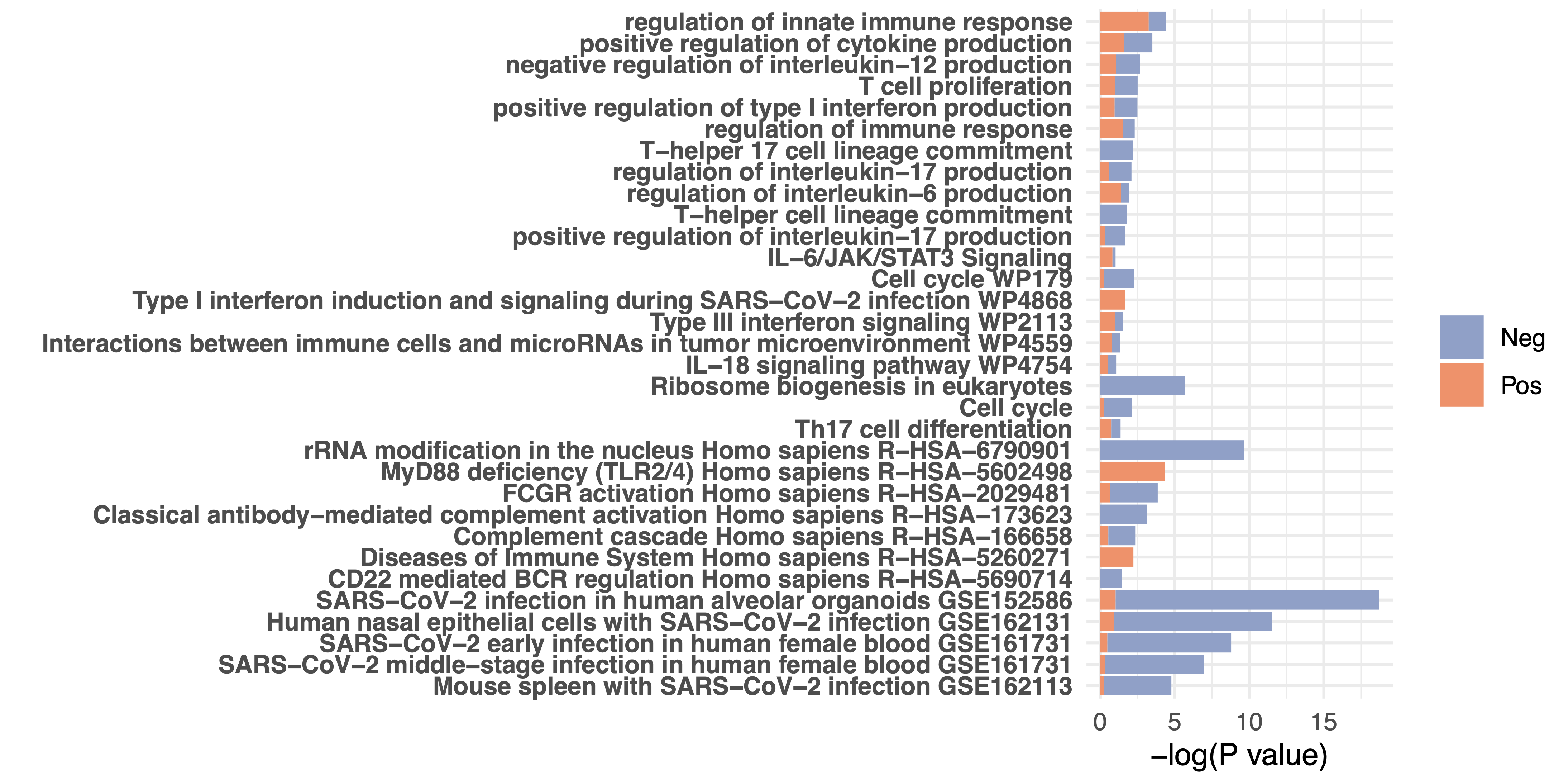

**Figure S2. Functional enrichment of COVID-19 severity associated genes in the 16 acute KTRs with T cell count information.** Differentially expressed genes (DEGs) in KTRs with high severity score (6-7) versus moderate severity score (3-5) were derived at nominal p-value ≤ 0.05, adjusted by age, gender and T cell count.

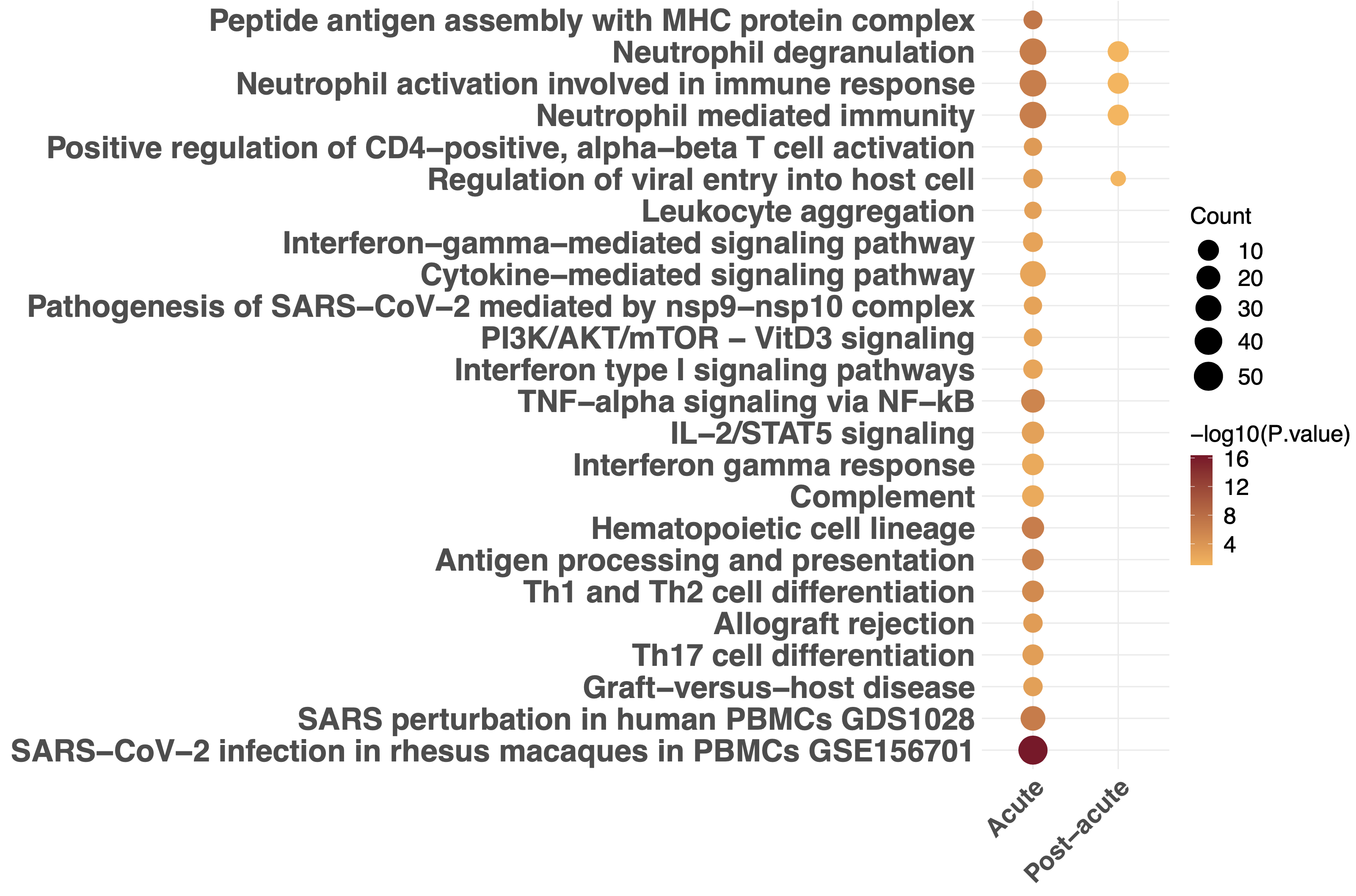

**Figure S3. Pathways enriched in DEGs associated with COVID-19 severity in acute and post-acute KTRs.** DEGs associated with the severity score were determined at nominal p-value ≤ 0.01 in acute and post-acute KTRs respectively. In acute KTRs, differential expression analysis was adjusted by age, gender, and lymphocyte count, while in post-acute KTRs, adjusted by age, gender, and days after initial COVID-19 diagnosis.

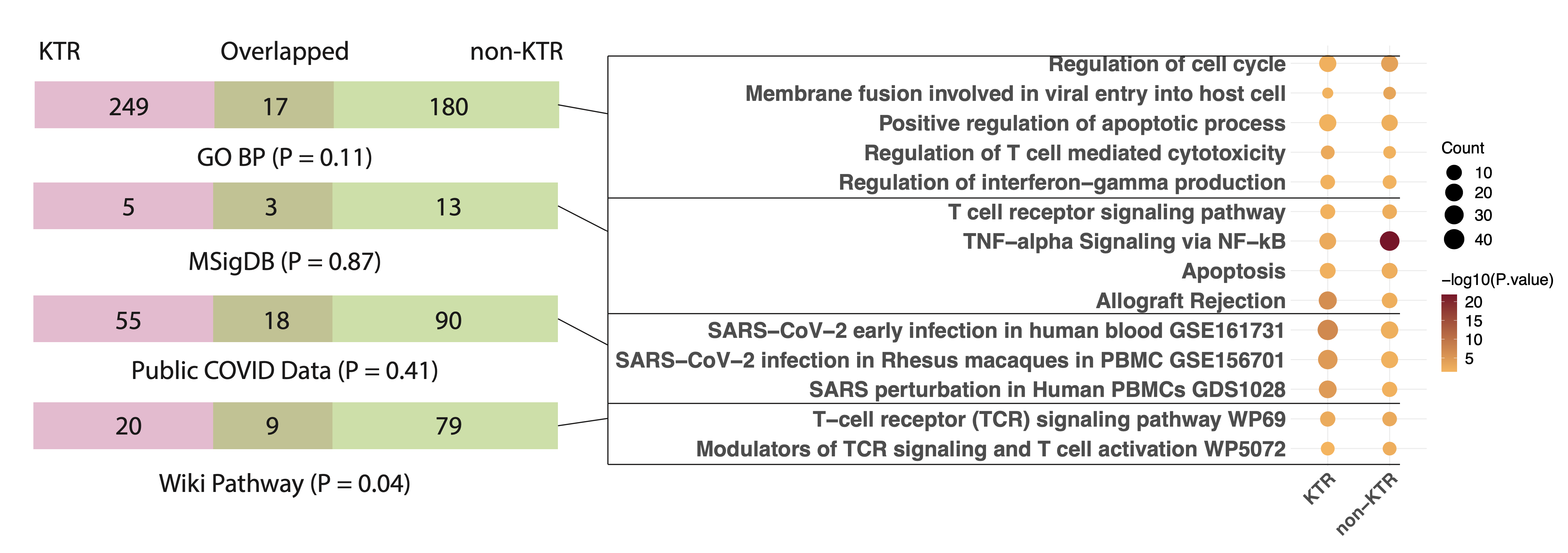

**Figure S4. Overlapped immune-related pathways in blood transcriptome of COVID-19 infected KTRs and non-KTRs.** Pathways are enriched in differentially expressed genes (DEGs) identified in post-acute patients as compared to acute patients. On the left panel, each Venn diagram (displayed in the form of bars) shows the overlap of enriched pathways between KTRs and non-KTRs (GEO accession: GSE157859).(*1*) The overlap significance was evaluated with a hypergeometric test. On the right panel, the representative overlapped pathways were listed in the dot plot with the size of the dot indicating the number of DEGs in each pathway and the color indicating the enrichment significance.

**Supplementary Tables**

**Table S1. Demographic and clinicopathologic characteristics of the acute KTRs stratified by the COVID-19 severity.**

| **Variable**^a^ | **Low**^b^ (n =8) | **Median**^b^ (n = 10) | **High**^b^ (n = 13) | **P-value**^c^ |
| --- | --- | --- | --- | --- |
| **Demographics** |  |  |  |  |
| Age (years) | 54 ± 13.44 | 46 ± 16.21 | 62.62 ± 10.09 | **0.02** |
| Gender, male | 5 (62.5) | 6 (60) | 5 (38.5) | 0.48 |
| Race, African American | 2 (25) | 4 (40) | 4 (30.8) | 0.79 |
| Body mass index (BMI) (kg/m^2^) | 30.55 ± 3.54 | 31.46 ± 4.8 | 29.47 ± 6.24 | 0.66 |
| Time from COVID-19 diagnosis (days) | 9.38 ± 8.62 | 7 ± 5.1 | 10.54 ± 6.98 | 0.48 |
| **Transplant information** |  |  |  |  |
| Transplant vintage (years) | 3.3 ± 3.6 | 7.6 ± 7.8 | 7.9 ± 7.4 | 0.39 |
| Donor status, living donor | 4 (50) | 4 (40) | 7 (53.8) | 0.90 |
| T-cell depletion induction | 4 (50) | 8 (80) | 6 (46.2) | 0.11 |
| Immunosuppression modification |  |  |  |  |
| Calcineurin inhibitors (CNI) |  |  |  | 0.22 |
| Not on CNI | 0 (0) | 1 (10) | 1 (7.7) |  |
| >50% reduction in CNI | 4 (50) | 3 (30) | 5 (38.5) |  |
| <50% reduction in CNI | 2 (25) | 6 (60) | 7 (53.8) |  |
| No change in CNI | 2 (25) | 0 (0) | 0 (0) |  |
| Mycophenolate Mofetil (MMF) |  |  |  | 1.00 |
| Not on MMF | 0 (0) | 0 (0) | 2 (15.4) |  |
| Similar dose | 0 (0) | 0 (0) | 0 (0) |  |
| >50% reduction or off | 4 (50) | 4 (40) | 5 (38.5) |  |
| Held dose | 4 (50) | 6 (60) | 6 (46.2) |  |
| Steroids (in Prednisone equivalents) |  |  |  | **0.02** |
| <0.5 mg | 8 (100) | 8 (80) | 5 (38.5) |  |
| 0.5 ~ 1 mg | 0 (0) | 2 (20) | 3 (23.1) |  |
| >1 mg | 0 (0) | 0 (0) | 5 (38.5) |  |
| **Comorbidity** |  |  |  |  |
| Smoking | 2 (25) | 1 (10) | 0 (0) | 0.10 |
| Diabetes | 5 (62.5) | 6 (60) | 7 (53.8) | 1.00 |
| Hypertension | 7 (87.5) | 10 (100) | 12 (92.3) | 0.72 |
| ACEI/ARB^d^ | 1 (12.5) | 1 (10) | 4 (30.8) | 0.52 |
| **COVID-19 symptoms** |  |  |  |  |
| Respiratory symptoms | 2 (25) | 7 (70) | 9 (69.2) | 0.09 |
| **Transplant outcome** |  |  |  |  |
| Acute kidney injury | 4 (50) | 10 (100) | 13 (100) | 1.00 |
| Graft loss within one year | 0 (0) | 0 (0) | 1 (7.7) | 1.00 |
| Death | 0 (0) | 0 (0) | 8 (61.5) | **<0.01** |
| Lymphocytes (10^3^/ml) | 1.46 ± 0.7 | 0.5 ± 0.24 | 0.62 ± 0.4 | **<0.01** |
| Baseline serum Creatinine (mg/dl) | 1.07 ± 0.23 | 2.76 ± 2.41 | 1.3 ± 0.44 | **0.03** |
| Peak serum Creatinine (mg/dl) | 2.34 ± 2.34 | 6.19 ± 6.11 | 3.1 ± 1.8 | 0.12 |

^a^: Unless otherwise specified, numeric variables are summarized as mean ± SD, and categorical variables as counts (percentage).

^b^: Low: COVID-19 severity score 1-3; Median: COVID-19 severity score 4-5; High: COVID-19 severity score 6-7.

^c^: In comparing between different severity strata, p-values were calculated by ANOVA for numeric variables and Fisher’s Exact test or Chi-square test for categorical variables. Bold p-value < 0.05.

^d^: ACEI/ARB: Angiotensin-converting enzyme inhibitor/Angiotensin receptor blocker.

**Table S2. Pathways enriched in DEGs associated with COVID-19 severity in acute KTRs.**

(Supplementary_Tables_S2_to_S7.xlsx)

**Table S3. Overlapped pathways enriched in DEGs associated with COVID-19 severity in acute KTRs and non-KTRs.**

(Supplementary_Tables_S2_to_S7.xlsx)

**Table S4. Pathways enriched in DEGs between post-acute versus acute KTRs.**

(Supplementary_Tables_S2_to_S7.xlsx)

**Table S5. Overlap of pathways enriched in DEGs associated with COVID-19 severity in acute KTRs and in DEGs between post-acute versus acute KTRs.**

(Supplementary_Tables_S2_to_S7.xlsx)

**Table S6. Neutrophil related functions are not enriched in DEGs between post-acute versus acute KTRs.**

(Supplementary_Tables_S2_to_S7.xlsx)

**Table S7. Overlapped pathways enriched in DEGs between post-acute versus acute in KTRs and non-KTRs.**

(Supplementary_Tables_S2_to_S7.xlsx)

**Table S8. Flow cytometry analysis on total blood or Ficoll-isolated PBMCs in a subset of acute KTRs.**

| **Subset** | **Markers** | **Parent population** |
| --- | --- | --- |
| Total lymphocytes | - | Acquired cells |
| Total T cells | CD3+ | Acquired cells |
| CD4+ T cells | CD3+CD8-CD4+ | CD3+ cells |
| CD8+ T cells | CD3+CD4-CD8+ | CD3+ cells |
